# Navigated ultrasound bronchoscopy with integrated positron emission tomography - A human feasibility study

**DOI:** 10.1101/2024.06.18.24308570

**Authors:** Arne Kildahl-Andersen, Erlend Fagertun Hofstad, Ole-Vegard Solberg, Hanne Sorger, Tore Amundsen, Thomas Langø, Håkon Olav Leira

## Abstract

**Background and objective:** Patients suspected to have lung cancer, undergo endobronchial ultrasound bronchoscopy (EBUS) for the purpose of diagnosis and staging. For presumptive curable patients, the EBUS bronchoscopy is planned based on images and data from computed tomography (CT) images and positron emission tomography (PET). Our study aimed to evaluate the feasibility of a multimodal electromagnetic navigation platform for EBUS bronchoscopy, integrating ultrasound and segmented CT, and PET scan imaging data.

**Methods:** The proof-of-concept study included patients with suspected lung cancer and pathological mediastinal/hilar lymph nodes identified on both CT and PET scans. Images obtained from these two modalities were segmented to delineate target lymph nodes and then incorporated into the CustusX navigation platform. The EBUS bronchoscope was equipped with a sensor, calibrated, and affixed to a 3D printed click-on device positioned at the bronchoscope’s tip. Navigation accuracy was measured postoperatively using ultrasound recordings.

**Results:** The study enrolled three patients, all presenting with suspected mediastinal lymph node metastasis (N1-3). All PET-positive lymph nodes were displayed in the navigation platform during the EBUS procedures. In total, five distinct lymph nodes were sampled, yielding malignant cells from three nodes and lymphocytes from the remaining two. The median accuracy of the navigation system was 7.7 mm.

**Conclusion:** Our study introduces a feasible multimodal electromagnetic navigation platform that combines intraoperative ultrasound with preoperative segmented CT and PET imaging data for EBUS lymph node staging examinations. This innovative approach holds promise for enhancing the accuracy and effectiveness of EBUS procedures.

## Introduction

Lung cancer stands as a leading cause of cancer-related death worldwide with a 18% of all cancer-fatalities in 2020 [1]. Early detection and efficient work-up is crucial for a favourable prognosis, as lung cancer is curable at an early stage. To identify patients eligible for curative treatment imaging with computed tomography (CT) followed by 18F-fluorodeoxyglucose (FDG) positron emission tomography (PET) scan is needed. These examinations form the basis for a clinical TNM staging, which describes size and features of the primary tumor (T), as well as involvement of hilar and mediastinal nodes (N) and the presence of distant metastasis (M). Subsequently, the radiological findings must be confirmed by invasive sampling, providing both a diagnosis and accurate staging information.

Endobronchial ultrasound bronchoscopy (EBUS) is the state-of-the-art tissue examination of PET-positive and/or enlarged lymph nodes (N-stage) within the lung hilus and mediastinum. Moreover, comprehensive mediastinal and hilar EBUS screening is strongly advised in scenarios involving tumors larger than 3 cm, central tumors, and cases with radiological N1-involvement [2]. This recommendation stems from the recognition that PET-CT scans, especially those indicating only hilar lymph node involvement (N1) or demonstrating a negative lymph node status (N0), may occasionally yield false negatives. An EBUS upstaging of 6% is shown in an observational study of Naur et al in such cases [3].

Bronchoscope with convex EBUS (CP-EBUS) probe enables real-time visualized or guided transbronchial needle aspirations (TBNA). Current lung cancer guidelines recommend sampling lymph nodes down to 5 mm in short axis when treatment to cure is intended [4]. At least three repeated needle aspirations are recommended for diagnosing lung cancer without rapid onsite evaluations (ROSE), even more to ensure an ample quantity of material for subsequent immunohistochemical and mutational analysis [4]. Notably, a study by Lee et al demonstrates that three needle aspirations exhibit a negative predictive value of 97%, proving effective in ruling out malignant involvement of a lymph node [5]. Assessment of hilar or mediastinal lymph node in lung cancer work-up with EBUS TBNA has shown specificity and sensitivity > 90% in meta-analysis [6, 7]. Additional procedures like elastography and ROSE have not contributed to higher sensitivity [8].

Limitations of EBUS include dependence on operator experience and individual learning curves, as evident in several studies [9–11]. The bronchoscopist must comprehend the mediastinal anatomy and possess a well-trained ability to mentally visualize preprocedural CT scans in three dimensions. To locate a specific lymph node station the initial navigation is mainly video-dependent. Subsequently, ultrasound localization of the lymph node(s) follows, presenting challenges such as a limited 60° ultrasound scan sector and occasional suboptimal images due to ultrasound artifacts and variable probe-to-wall contact. Furthermore, the ability to perform repeated aspirations on the same lymph node is crucial for obtaining representative samples. However, lymph node relocation becomes challenging after each aspiration, as the video view is often obscured by blood and mucus.

Electromagnetic navigation (EM) bronchoscopy (ENB) was first demonstrated successfully in humans in a study of Schwarz et al in 2006 [12]. Since then, ENB has been a growing field of research for diagnosing lung lesions located beyond visual access in the periphery of the lungs. However, Sorger et al has presented a system for ENB suitable for diagnosing mediastinal/hilar lymph nodes accessible for CP-EBUS [13, 14]. The clinical rationale behind incorporating ENB with EBUS lies in addressing the challenges associated with video dependency. In this study, the tracking sensor were permanently fixated to the EBUS bronchoscope with glue and endoscopy rubber wrap. Not confined to one EBUS bronchoscope, a single-use 3D printed click-on device to attach the EM tracking sensor was developed [15]. However, this click-on device has never been tested with ENB in patients.

Our study introduces novel features as the fixation click-on device for EM tracking sensor including a preoperative tracking sensor-to-ultrasound image calibration procedure. Methods for CT segmentations are implemented and a real-time integration of PET-CT data during EBUS bronchoscopy is introduced, allowing for the identification of PET-positive lymph nodes during ENB. Our study aimed to evaluate the feasibility of a complete multimodal image guidance platform for navigated CP-EBUS in humans.

## Material and methods

### Study design and patients

The study was designed as a non-randomized proof-of-concept study with patients recruited prospectively. Inclusion criteria were patients > 18 years, non-pregnant, and referred to the thoracic department with suspected lung cancer and possible curable disease. Patients eligible for participation exhibited pathological mediastinal or hilar lymph nodes warranting a PET-CT examination. The study was approved by the regional ethics committee (ID 13136), being in accordance with the Declaration of Helsinki. Written and informed consent was collected from the patients. The patients were included from August to October 2023.

### Electromagnetic navigation platform

The ENB system is constructed upon the open-source navigation research platform, CustusX [16]. The preoperative and intraoperative setup steps of the ENB system are illustrated in Figure 1. Preoperative radiologic imaging data underwent automated airway segmentation processes [17], and artificial intelligence (AI)-based segmentation of lymph nodes and vessels [18]. Image-to-patient registration was performed by automated airway centerline estimations of the tracked EBUS bronchoscopés positions during the initial phase of the procedure [19]. The tracking sensor was affixed to the distal end of the EBUS bronchoscope using a 3D printed clip-on device made of Ultem™ 1010 resin developed in our research group. The disposable clip-on device has been tested previously on a healthy volunteer [15]. A quick on-site EM tracking-to-ultrasound calibration was performed using a probe calibration adapter [15]. During the bronchoscopy, CustusX displays a combination of preoperative and real-time intraoperative images. A graphical user interface with both video, ultrasound, virtual bronchoscopy, or CT could then be obtained. Finally, by combining different image modalities with EM tracking a navigation map is presented for the bronchoscopist in real-time to guide the procedure.

**Fig 1.**
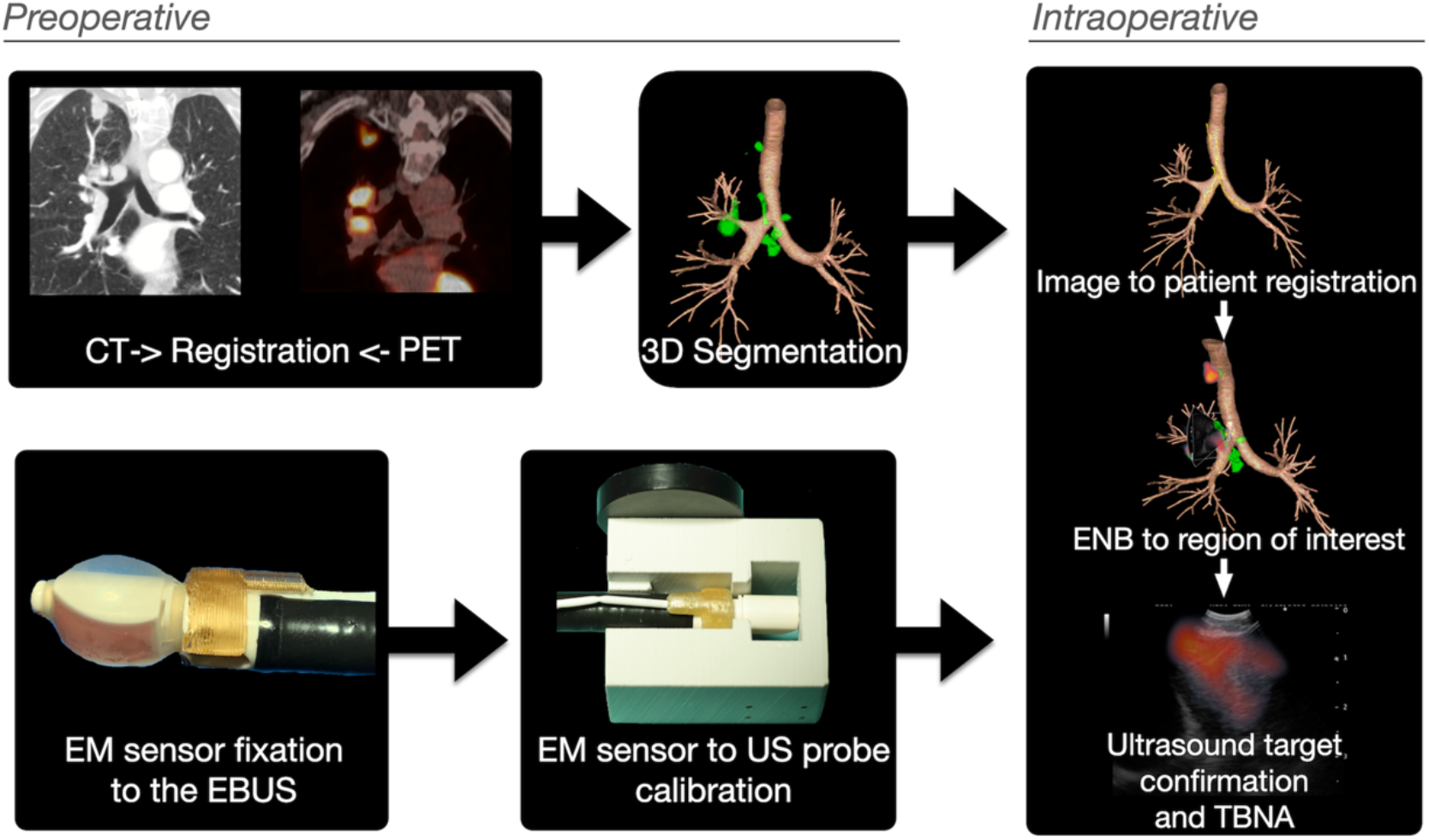
The steps of image preprocessing and EBUS bronchoscope modifications that enables electromagnetic navigation bronchoscopy (ENB). Airways and lymph nodes are segmented from the CT images and registered to PET. An electromagnetic (EM) sensor is fixated to the EBUS bronchoscope and calibrated to the ultrasound scan sector with a 3D-printed adapter. Intraoperatively the CT/PET images are registered to the patient, before enabling the bronchoscopist to navigate to the target supported by ENB and ultrasound

### Equipment for bronchoscopy electromagnetic position tracking system

- CP-EBUS (Olympus BF-UC180F, Olympus, Tokyo, Japan)
- Video processing unit (Olympus, Evis Excera III CV-190 Plus, Olympus, Tokyo, Japan)
- Light source (Olympus, Evis Excera III CLV-190, Olympus, Tokyo, Japan)
- Ultrasound processor (Olympus EVIS EUS EU-ME2, Olympus, Tokyo, Japan)
- CustusX (In-house developed open-source research system for image guiding and navigation, www.custusx.org) [16]
- Aurora^®^ electromagnetic tracking system (Northern Digital Inc. (NDI), Waterloo, ON, Canada)
- Tracking sensor (Aurora 6-DOF Probe, Straight Tip, Standard, Northern Digital Inc., Waterloo, ON, Canada)
- 3D printed clip-on device made in Ultem™ for attaching the electromagnetic tracking sensor to the bronchoscope [15]
- 3D printed EBUS bronchoscope adapter for calibration [15]

### Integrating PET-CT imaging data into the navigation platform

Patients included in the study underwent a PET-CT scan as part of the staging process for suspected lung cancer. However, the lowdose CT scan acquired simultaneously to merge with the PET tracer imaging data has limited resolution and is without intravenous contrast. To enhance the precision of the imaging, a high-resolution thorax CT scan with a slice thickness of 1 mm, conducted weeks before the PET examination, was registered to the lowdose PET-CT data. This registration process involved a 3-D deformable registration, necessary as the PET imaging data was captured during different respiratory phases compared to the initial high-resolution CT. The Elastix toolbox was integrated into CustusX for this purpose [20, 21].

### Technical and clinical outcomes

Navigation accuracies were measured postoperatively. This involved a manual operation using both lymph nodes and large vessels displayed on the ultrasound recordings as gold standard. The positions of these structures were then compared to their counterparts in the segmented 3-D CT map. The total procedure time of the EBUS examinations was recorded. The TBNA were carried out without deviation from routine clinical practice, where the primary outcome was to successfully achieve a diagnosis from each sampled lymph node. The 3D printed clip-on device with EM sensor was qualitatively evaluated by inspection after each procedure to determine if it remained robustly attached to the CP-EBUS.

## Results

Included in the study were two female and one male referred to the department of thoracic medicine due to symptoms of suspected lung cancer. The CT and PET imaging data were segmented and displayed intraoperatively. Segmented images rendered on the navigation platform are shown alongside ultrasound in Figs 2 to 4. All lymph nodes were correctly segmented, with the exception of one PET-negative station 7 node from patient 1. The timespan separating high-resolution CT and PET-CT to EBUS ranged from 16 to 29 days and 6 to 10 days, respectively. After PET-CT patient 3 had questionable distant lymph node spread to left thigh (not puncturable). The TNM-stage of the patients is shown in Table 1.

**Fig 2.**
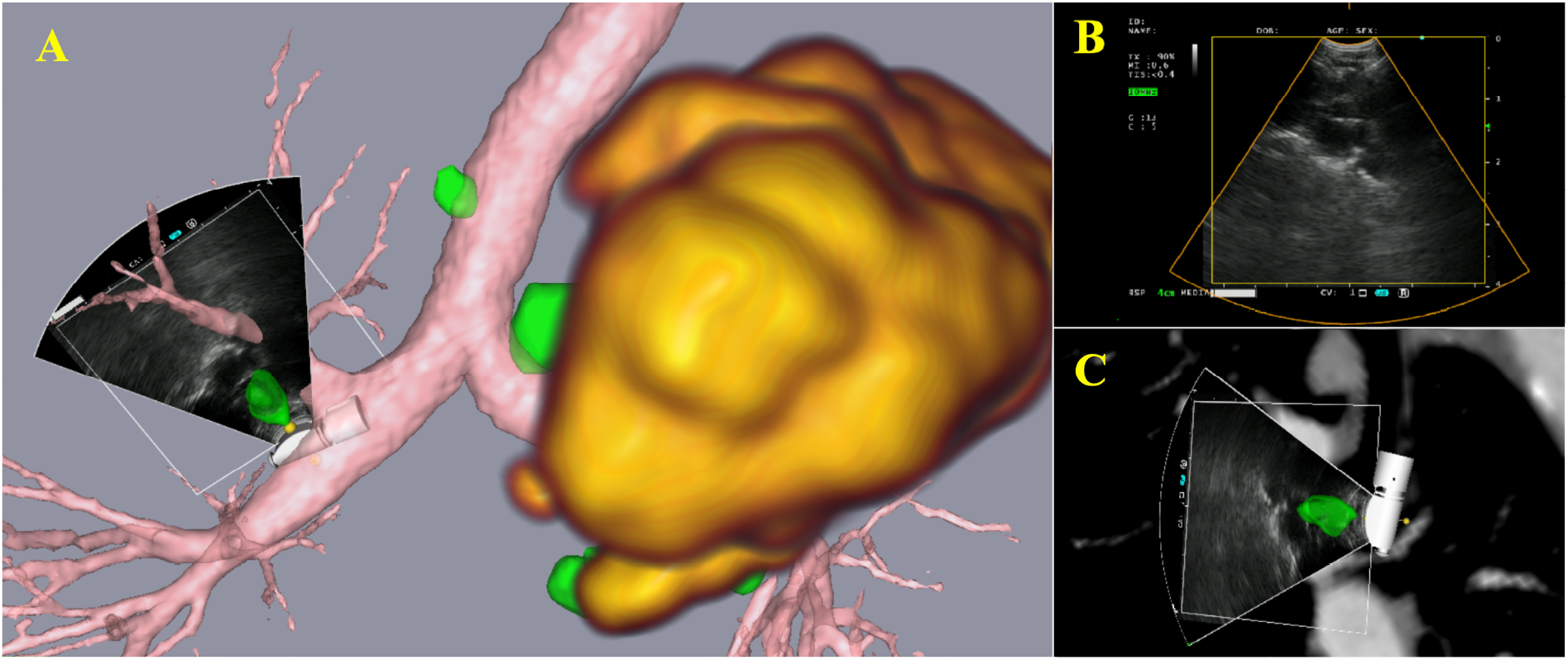
Graphical renderings from patient 1 as displayed on the navigation platform. In the 3D renderings on the left (A), the ultrasound scan sector showing lymph node station 11R. A lung tumor in the upper lobe of the left lung is highlighted in yellow, indicative of PET positivity, while the lymph nodes are depicted in green, signifying PET negativity. The top right (B) displays the ultrasound view from the EBUS bronchoscope, and in the lower right (C), the EBUS bronchoscope’s probe and ultrasound scan sector is visualized together with the CT images and overlaid the segmented lymph node (green).

**Fig 3.**
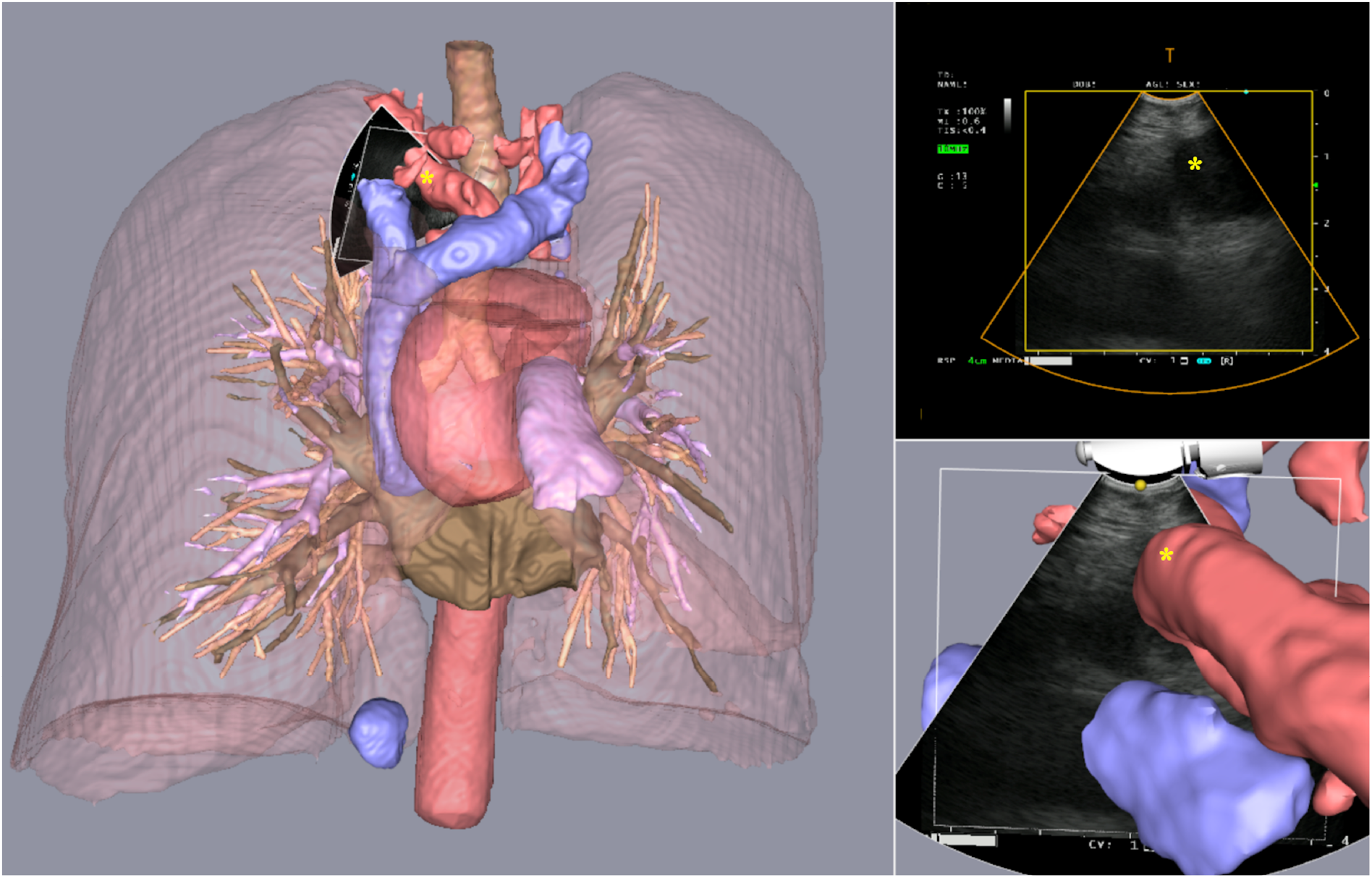
Segmented 3D CT renderings, patient 2. Great thoracic vessels displayed intraoperatively, with the ultrasound scan sector showing the left subclavian artery (*).

**Fig 4.**
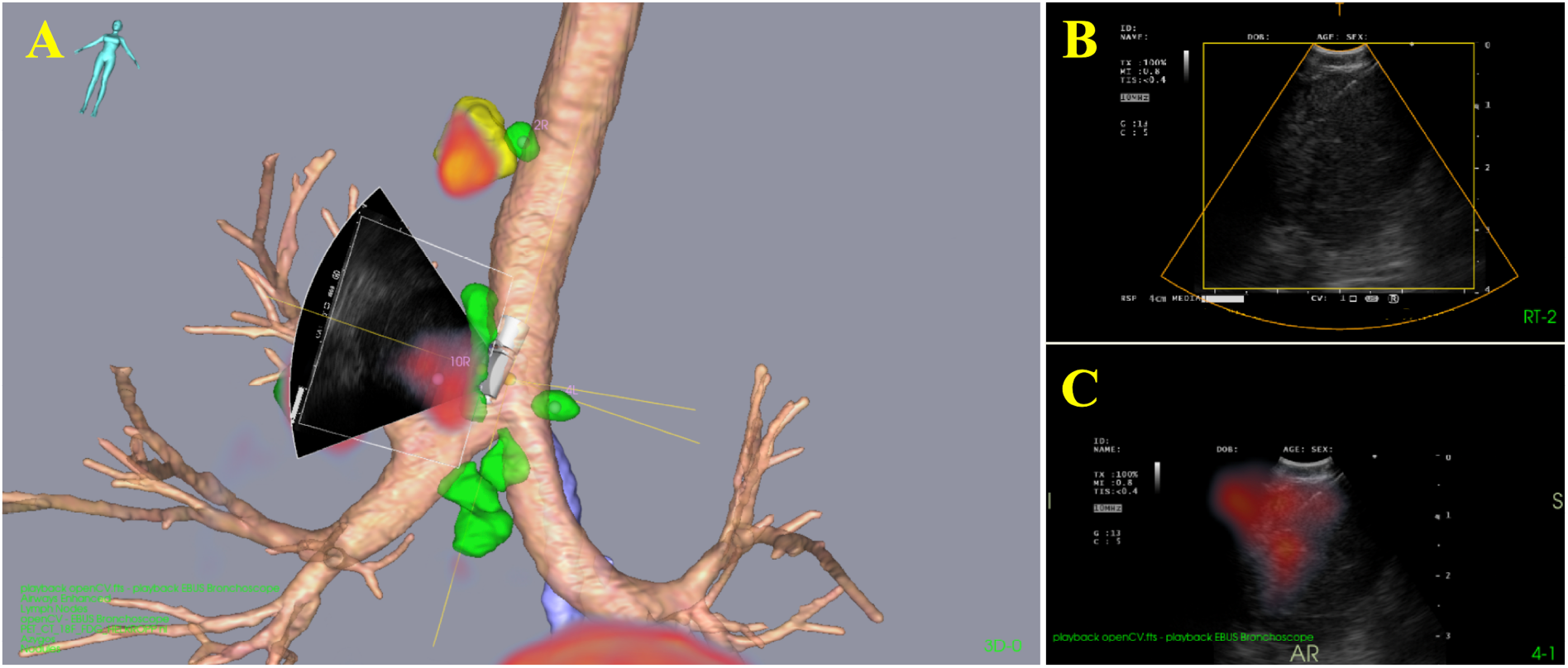
Graphical user interface during sampling, patient 3. In the segmented 3D CT and PET-CT renderings (A), the EBUS probe is shown during needle aspirations of lymph node station 10R. The top right (B) illustrates the ultrasound view from the bronchoscope, showing both the needle and a relatively large lymph node. The lymph node, identified as PET-positive, thereby displayed in red in both the hybrid PET-CT-ultrasound image (C) and the 3D PET-CT renderings (A). The EBUS bronchoscopy procedures lasted between 20 to 25 minutes for all three patients. Except for the initial registration of the preoperative CT data to the patient, procedures were not impeded by the additional equipment. Rapid onsite examination (ROSE) was only employed for patient 3. Malignant cells were found in the sampled lymph node stations in two of the three patients. In the first patient, EBUS sampling results revealed lymphoid cells, but squamous cell carcinoma (SCC) was diagnosed with four forceps biopsies from the primary tumor. Detailed results from imaging and EBUS examinations are provided in Table 1.

**Table 1.** Three patients examined with navigated EBUS: Technical outcomes

| Patient | Stage (TNM) | Primary tumor<br>size (mm) | LN stations | Size<sup>§</sup><br>(mm) | Median<br>accuracy<br>(mm) | Range<br>(mm) | Aspirations | Diagnosis | Duration<br>(min) |
| --- | --- | --- | --- | --- | --- | --- | --- | --- | --- |
| **1** | cT4N2M0 | 114 | 7 | 9 | NA | NA | 3 | LC | 20 |
|  |  |  | 11R | 10 | 8.3 | 3.9-13.3 | 4 | LC |  |
| **2** | cT4N3M0 | 89 | 2R | 16 | 8.3 | 7.9-10.5 | 4 | NSCLC | 23 |
|  |  |  | 4R | 8 | 7.0 | 6.7-7.2 | 4 | NSCLC |  |
| **3** | cT1cN1(3)M1b | 18 | 10R | 20 | 3.5 | 3.1-6.8 | 5 | NSCLC | 25 |

The table shows sampled lymph node (LN) stations, size (from CT scan), median accuracy for each station and number of needle
aspirations(passes). In addition to a NSCLC diagnosis, the provided cytologies from patient 2 showed anaplastic lymphoma kinase (ALK)
positive cancer.
§ CT scan lymph node size in short axis
LC= Lymphoid cells, NSCLC=Non-small cell lung cancer

The 3D printed clip-on device affixed at the tip of the EBUS bronchoscopy remained securely attached throughout the examinations. Importantly, all bronchoscopies were performed without complications for the patients, demonstrating no excess bleeding, pneumothorax, or post-procedure infections.

The navigation accuracies for the three patients were measured postoperatively and were assessed by analyzing image recordings from the navigation platform. A total of 24 target points, sampled from ten distinct anatomical locations, were included in the measurements. Given that ultrasound 3D acquisitions were beyond the scope of this study, the measurements were confined to 2D ultrasound images. To mitigate bias, measurements were systematically taken from various scan sector directions.

Median accuracy estimated from the ten anatomical positions was 7.7 mm, and range was 3.3-15.3 mm.

## Discussion

This proof-of-concept study establishes the feasibility of a multimodal ENB platform for the staging of mediastinal and hilar lymph nodes in lung cancer. The platform integrates multimodal and elastic co-registration of images (CT and PET), EBUS, and AI-based segmentation of central airways, vessels, and lymph node. The final result is a multimode ENB, PET CT, and EBUS guiding platform for the mediastinal/hilar lymph nodes. By adding PET-CT to the ENB platform the bronchoscopist is able to discern between PET positive and negative lymph nodes in real-time. This is a useful feature to guide sampling, especially in cases where PET positive and negative lymph nodes coexist in the same lymph node station.

The median accuracy of 7.7 mm compares well with findings from prior articles on similar subject [14, 22]. Notably, the measured accuracy appears reasonable when considering a preliminary phantom study involving the current click-on device and preprocedural calibration system, demonstrating an accuracy of 3.9 mm [15]. In a previous study utilizing the CustusX navigation platform, Sorger et al reported an overall accuracy of 10.0 mm in examining four patients [14]. Other studies utilizing ENB for peripheral lung tumors have indicated navigation accuracy ranging from 5.7 to 9.0 mm [22]. When utilizing ENB on humans, additional offset values are introduced due to cardiorespiratory movements and displacement of lung tissue by the ultrasound probe pushing the bronchial wall [23]. Our study indicates a greater impact on navigation accuracy errors in the hilus region, specifically at station 11, as this area is more susceptible to respiratory movements and probe induced displacements than stations situated along the trachea [23]. This accuracy error becomes more clinically relevant in the clinical context of transbronchial sampling of peripheral lung tumors without real-time ultrasound guidance.

A virtual EBUS bronchoscopy that enables preoperative planning and intraoperative guidance of bronchial wall puncture sites was presented in a study of Sato et al [24]. A virtual EBUS was performed first to determine puncture spots and angels, followed by a real EBUS examination of the patient performed based upon this information. In contrast, our system offers instantaneous EBUS to CT localization within the airways, presenting a valuable advantage. Addressing a practical concern with traditional EBUS bronchoscopy, where mucus or blood can obscure the bronchoscope video during needle aspirations, EM navigation virtual bronchoscopy (from 3D CT), can be used independent from the real video. Ensuring repeated needle aspirations come from the same lymph node is crucial for bronchoscopists. ENB facilitates swift relocation to the same lymph node when repeated aspirations are necessary, potentially optimizing procedure duration. Finally, adding ENB to EBUS will grant navigational assurance for the novice bronchoscopists unfamiliar to EBUS, as it displays the bronchoscope’s anatomical 3D position relative to the mediastinal and hilar anatomy.

The studies of Zang et al outline a system combining virtual video bronchoscopy and CT-based virtual EBUS registered to real EBUS [25, 26]. The proposed method combines segmentation of both ultrasound and CT images that are fused through a six-step algorithm based on mutual information and region shape information. The method has advantages such as not requiring extra sensors or navigation hardware, but segmentation of nodal regions of interest (ROI) was time-consuming. As often is the case, many lymph nodes are to be examined, so that total segmentation time could be perceived disruptive in a EBUS workflow, especially when patients are under conscious sedation. Despite this, both navigation systems, including ours, prove beneficial for learning purposes for novice EBUS bronchoscopists, addressing the challenging learning curve associated with EBUS.

Compared to a software-only solution, a complete EM platform setup with calibration and registration to patients is more complex and time consuming. Inherent navigation accuracy errors, metallic hardware interference, registration errors, and cardiorespiratory motions contribute to CT-to-body divergence. However, ENB systems have demonstrated efficacy in diagnosing peripheral lung nodules [27]. In clinical scenarios with both peripheral tumors and enlarged mediastinal lymph nodes, using ENB for navigation to both sites during the same session is justifiable. Nonetheless, ENB competes with alternative techniques, like tomosynthesis and robotics [28], and its future role, especially in the lung periphery, remains to be determined [29].

Our study limitations include the small number of participants and the introduction of a 3D printed clip-on device, which, though complication-free in our study, could pose potential issues. Future considerations may involve tracking cytology needles electromagnetically, eliminating the need for externally attached sensors. Segmentation of one of the lymph nodes failed, showing that manual verification before bronchoscopy remains essential. Finally, the postprocedural measurements were limited to only two orthogonal planes in the 2D ultrasound scan sector. Though, by measuring accuracy from different anatomical sites and ultrasound scan directions we aimed at lowering the likelihood of bias in the navigation accuracy measurements.

Diagnostic yield and procedural time reduction could be potential benefits of navigation platforms, which could be explored in future trials. The intraoperative captured multimodal images could serve as quality indicators and documentations that could be reviewed on multi-disciplinary meetings. Our ongoing research explores machine learning for recognizing specific lymph node stations during EBUS bronchoscopy, with ENB serving as a golden standard linking patient anatomy from CT to intraoperative video and ultrasound recordings.

In conclusion, our study demonstrates the feasibility of integrating PET imaging data into a multimodal EM navigation platform for lymph node staging examinations. Introducing PET data during real-time EBUS provides valuable information for prioritizing invasive sampling. Future trials and advancements, including further image analytics with machine learning, will unveil the full potential of navigation platforms in enhancing bronchoscopy procedures.

## Data Availability

Ultrasound, CT and PET data from the study will be made available online, with link for download in the paper, upon publication acceptance.

## Acknowledgements

We thank Marit Setvik and Solfrid Meier Løwensprung, Department of Thoracic Medicine, St. Olavs hospital, for their contribution in assisting the bronchoscopy. This work was supported by St. Olavs hospital, the Liaison Committee for Education, Research and Innovation in Central Norway and SINTEF Research group for Medical Technology, the Ministry of Health, and Social Affairs of Norway through the Norwegian National Advisory Unit for Ultrasound an Image-Guided Therapy (St. Olavs hospital, Trondheim, Norway)

